# Mechanistic models of humoral kinetics following COVID-19 vaccination

**DOI:** 10.1101/2024.02.08.24302502

**Authors:** Daniel Stocks, Amy Thomas, Adam Finn, Leon Danon, Ellen Brooks-Pollock

## Abstract

**Introduction:** Future COVID-19 vaccine programmes need to take into account the variable responses elicited by different vaccines and their waning protection over time. Existing descriptions of antibody response to COVID-19 vaccination convey limited information about the mechanisms of antibody production and maintenance.

**Methods:** We describe the antibody dynamics elicited by COVID-19 vaccination with two biologically-motivated mathematical models of antibody production by plasma cells and subsequent decay. We fit the models using Markov Chain Monte Carlo to seroprevalence data from 14,602 uninfected individuals collected via the primary care network in England between May 2020 and September 2022. We ensure our models are structurally and practically identifiable when using antibody data alone. We analyse the effect of age, vaccine type, number of doses, and the interval between doses on antibody production and longevity of response.

**Results:** We find evidence that individuals over 35 years of age who received a second dose of ChAdOx1-S generate a persistent antibody response suggestive of long-lived plasma cell induction, while individuals that receive two doses of BNT162b2, or one dose of either vaccine do not. We also find that plasamblast productive capacity, the likely driver of short-term antibody responses, is greater in younger people than older people (≤ 4.5 fold change in point estimates), people vaccinated with two doses than people vaccinated with one dose (≤ 12 fold change), and people vaccinated with BNT162b2 than people vaccinated with ChAdOx1-S (≤ 440 fold change). The effect of age on antibody dynamics is more pronounced in people vaccinated with BNT162b2 than people vaccinated with ChAdOx1-S. We find the half-life of an antibody to be between 23 – 106 days.

**Conclusion:** Routinely-collected seroprevalence data are a valuable source of information for characterising within-host mechanisms of antibody production and persistence. Extended sampling and linking seroprevalence data to outcomes would allow for powerful conclusions about how humoral kinetics protect against disease.

## Introduction

Vaccination against COVID-19 is now routinely used to maintain levels of population immunity. It is important to understand how the immune response elicited by vaccination will change with time. The antibody response to vaccination varies with age (1; 2; 3; 4), peaking within a couple of weeks and subsequently waning over the course of six months (4; 5; 6). The mRNA Pfizer-BioNTech vaccine (BNT162b2) elicits a stronger antibody response than the adenoviral vector AstraZeneca vaccine (ChAdOx1-S) after the first dose and up to two weeks after the second dose (7), with the response to the second dose being more than an order of magnitude greater than the response to the first dose (7; 8; 9; 10).

Mathematical models can be used to help understand underlying mechanisms of the response, explain differences between age and vaccines, and predict future antibody levels. Previous modelling of SARS-CoV-2 specific antibody levels used statistical, single-phase exponential decay models often implemented through linear regressions (11; 12; 13; 14; 15; 16). These models convey limited mechanistic information because they are not biologically motivated and are constrained by the inherent assumptions in their structure.

Analysis of the immune response is challenging because there are many unobserved components. Quantification of serum antibody concentrations offers a viable window into the dynamics of the humoral system (17; 18; 19). Serum antibody concentrations can be quantified by taking blood samples, and mucosal antibodies can be quantified in saliva or other mucosal samples, while plasma cells that reside in the bone marrow (20; 21; 22; 23) and their precursors, B cells, that differentiate in lymph nodes are much harder to accurately quantify without invasive procedures. It is therefore of interest to see whether information about the underlying kinetics of antibody production can be extracted from data on antibody level alone.

Binding antibody titers have been correlated with COVID-19 vaccine efficacy (24; 25), suggesting antibody titers are important for preventing the development of serious disease and reinfection (26). Similarly, antibody levels on admission to hospital are inversely correlated with in-hospital mortality (27).

### Maintenance of a persistent immune response

Biological mechanisms that maintain the humoral response after exposure (natural infection or vaccination) have been investigated (28). However, hypotheses that persisting antibody levels are a result of memory B cells being continuously stimulated through chronic infection, re-exposure, persistent antigen, or bystander T cells are not consistent with observations. The re-exposure and bystander T cell hypotheses (28) rely on pathogens being endemic and therefore cannot explain persisting antibodies in the absence of local outbreaks (29; 30; 31). The persistent antigen hypothesis is based on evidence that follicular dendritic cells (FDCs) sequester antigen in later stages of infection and, over time, display it to memory B cells to encourage a continued humoral immune response in the absence of infection (32; 33; 34). However, the decay of antigen retained by FDCs in mice is too fast to explain stable antibody production (35) although, persistent antigen could be important early in the memory response.

Other hypotheses explain persistent antibody levels with independently regulated plasma cells, instead of memory B cells. It has been shown that plasma cell populations and antibody levels persist in the absence of memory B cells (29; 36; 37; 38; 39; 40). One hypothesis that links plasma cell populations to antibody levels is competition between newly produced plasmablasts and residing, long-lived plasma cells for niches in the bone marrow (20; 21; 22; 23; 41). However, this hypothesis predicts a faster decline in long-term antibody levels in older people, which is not observed (29). A different hypothesis (the ‘*imprinted lifespan*’ hypothesis) proposed in (28) is that plasma cells are imprinted with a short or a long lifespan upon differentiation from B cells. B cells that differentiate without interaction with CD4^+^ T cells (T cell independent) and are cross-linked by repetitive foreign antigen become short-lived plasma cells (plasmablasts) (28). Whereas, a lifespan of years or decades is thought to be achieved by B cells that are cross-linked by a non-repetitive foreign antigen and interact with CD4^+^ T cell (T cell dependent) (28). To acquire even longer lifespans (potentially as long as the person themselves) B cells must be cross-linked by repetitive foreign antigen and interact with CD4^+^ T cells (28).

Following a mathematical formulation of the ‘*imprinted lifespan*’ hypothesis proposed in (18) we analyse two mathematical models. One model includes long-lived plasma cells, the other does not. Figure 1 shows a qualitative description of the modelled humoral dynamics, similar to those observed in vivo (42). We impose the requirement that models are structurally identifiable and then ensure that they are practically identifiable, i.e. that observed outputs can only be generated by a single parameter set (a different parameter set would generate a different output) (43; 44; 45), and that the data contain enough information to inform the parameters.

**Fig. 1.**
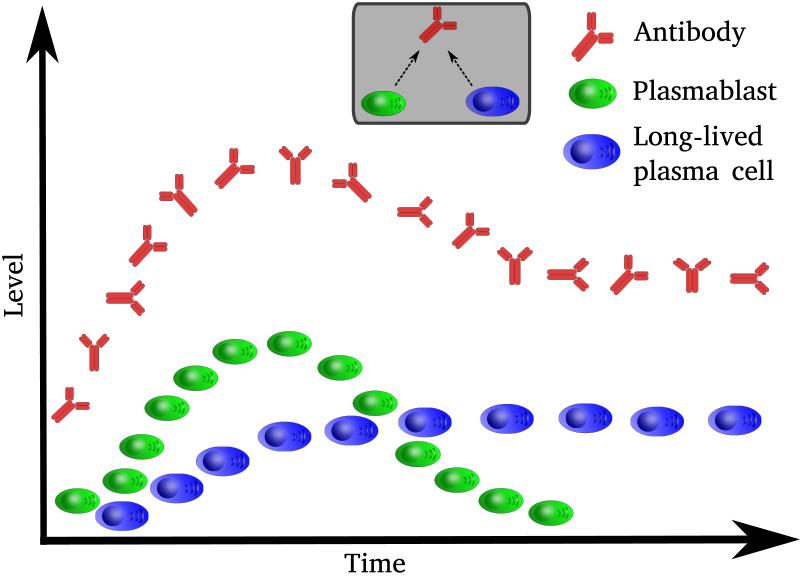
A schematic of modelled humoral dynamics. A persistent antibody response is dependent on the generation of long-lived plasma cells. In contrast, early antibody dynamics and peak serum antibody concentrations are driven by short-lived plasma cells (plasmablasts).

We assess parameter identifiability to understand which model describes the kinetics of the humoral response after one and two doses of COVID-19 vaccination for different ages and vaccine schedules. From our analysis, we draw conclusions on which groups show evidence of long-term antibody responses, how age and vaccine schedule impact antibody levels, and the importance of a second dose.

## Results

### Antibody dynamics by age and vaccine type

We split the data on antibody level into age groups (16 – 34, 35 – 54, 55 – 74, 75+), vaccine type (BNT162b2, ChAdOx1-S), and number of doses (one or two), prior to model fitting. Using two models of antibody dynamics, derived from biologically motivated equations of humoral kinetics (equation (2) and equation (3)), we are able to describe the SARS-CoV-2 specific antibody response of 7 of the 8 first dose groups and 8 of the 16 second dose groups, and make predictions about future antibody levels. Table 2 contains the groups we are able to model. A group can be modeled if using its data, all the parameters of one of the models are identifiable (i.e. the model is identifiable).

The antibody dynamics of all modelled first dose groups can be described by the single cell model, while the asymptotic model only describes 35 – 54 year-olds vaccinated with one dose of BNT162b2. However, the single cell model is a better predictor of the data collected from this group than the asymptotic model. Bayes’ Factor is 159,000 for the single cell model over the asymptotic model.

We are able to model seven of the second dose groups (see table 2). The antibody dynamics of 35 – 54 year-olds and 55 – 74 year-olds, and 75+ year-olds vaccinated with two doses of ChAdOx1-S is better described (measured by Bayes’ Factor) or can only be described by the asymptotic model. Data collected from 55 — 74 year-olds who received two doses of ChAdOx1-S greater than 77 days apart were better predicted by the asymptotic model, with a Bayes’ Factor of 63.0 for the asymptotic model over the single cell model. The antibody dynamics of 75+ year-olds vaccinated with two doses of BNT162b2 can only be described by the single cell model. Data collected from 16 — 34 year-olds who received two doses of BNT162b2 at most 77 days apart was better predicted by the asymptotic model, with a Bayes’ Factor of 12.1 for the asymptotic model over the single cell model.

Figure 2 shows the dynamics of the modelled groups given in table 2. For the first dose the models are initialised at zero (*A*_0_ = 0). It is assumed in the asymptotic and single cell models that the initial population of plasma cells is the full population that then decays. This assumption results in an immediate sharp rise in antibody levels. This sharp rise does not reflect what we observe in the data for the antibody response to the first dose but does for the response to the second dose.

**Fig. 2.**
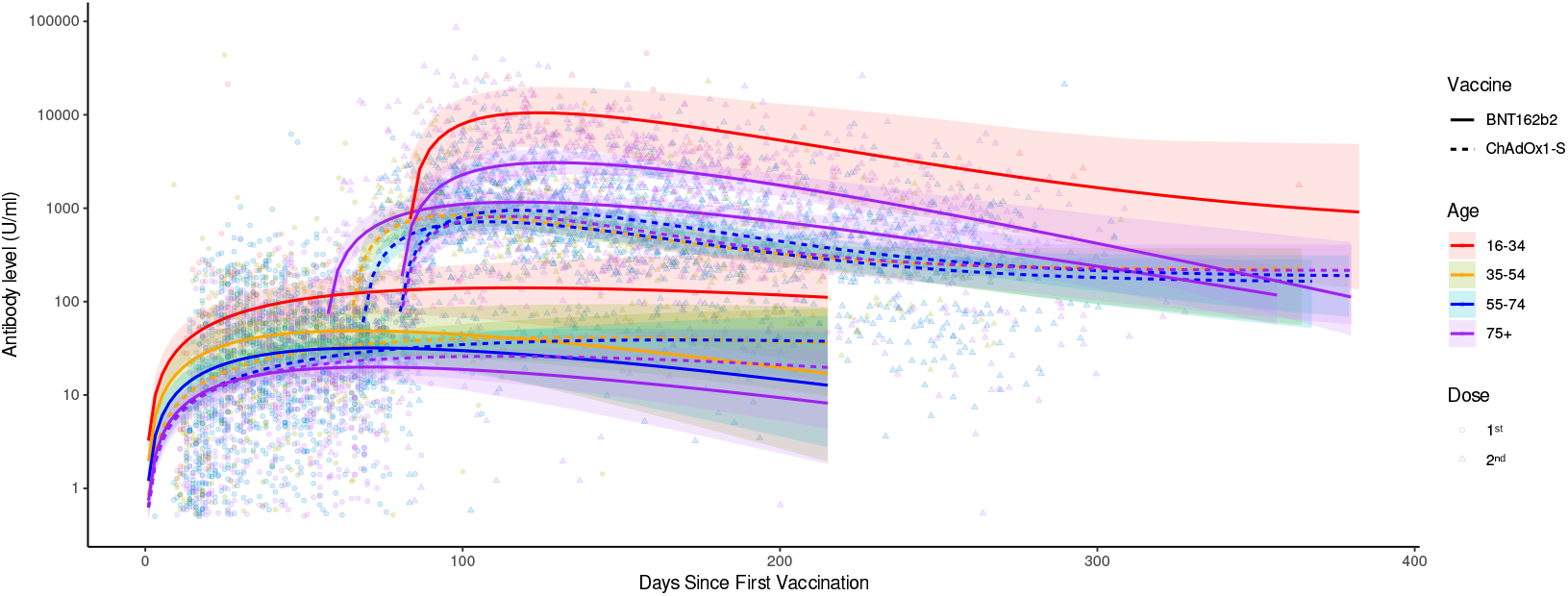
The antibody data and best fitting model of the group whose data could identify a model (the groups in table 2). The models for people who have received two doses ChAdOx1-S are the asymptotic model, and all others models are the single cell model. For the first dose we initialise the models at zero (*A*_0_ = 0). All parameter estimates are given in table 2. The shaded regions are defined by the 95% HPD CrI of the parameter estimates.

Age has a greater impact on the magnitude of the antibody response for people vaccinated with BNT162b2 than those vaccinated with ChAdOx1-S. After both the first and second dose, younger people vaccinated with BNT162b2 have higher antibody levels than older people vaccinated with BNT162b2 at all time points and our models predict this trend will continue into the future. While for those vaccinated with ChAdOx1-S there is little discrepancy between the age groups, especially after the second dose.

We also find that those vaccinated with one dose of BNT162b2 have higher antibody levels than the same age group vaccinated with one dose of ChAdOx1-S in the short-term but lower levels in the long-term. The length of time those vaccinated with BNT162b2 have higher antibody levels than the same age group vaccinated one dose of ChAdOx1-S is age dependent. Younger people vaccinated BNT162b2 have higher antibody levels than the same age group vaccinated with ChAdOx1-S for longer than older people. The time of cross-over is 113.4 days, 90.0 days, and 33.4 days, for 34 – 35 year-olds, 55 – 74 year-olds, and 75+ year-olds, respectively. However, the 95% HPD CrIs of the means overlap for the whole time-frame of the first dose data (215 days post vaccination), so we cannot say with confidence that a cross-over occurs in the timeframe of these data. We do not have a comparison for 16 – 34 year-olds because the data on this age group’s response to one dose of ChAdOx1-S were insufficient to identify a model.

We also find that people vaccinated with two doses of BNT162b2 will have higher antibody levels than those vaccinated with ChAdOx1-S for approximately nine months after receiving their second dose. However, our models predict that after nine months the levels will be similar and in the long-term, those vaccinated with ChAdOx1-S will have higher antibody levels. A possible exception is 16 – 34 year-olds vaccinated with BNT162b2. This group is best described by the asymptotic model. However, the 95% HDP CrI for the group’s antibody dynamics includes decaying to zero in the long-term. So, we cannot be confident 16 – 34 year-olds vaccinated with two doses of BNT162b2 will have persisting antibodies. This wide interval is probably due to the small sample size of the group (*n* = 25).

### Estimates of Humoral Response Parameters

Even though only 14 of the 24 groups we investigate can identify all the parameters of at least one of the proposed models and are therefore the only groups we can say the models describe, we can still extract information about individual, identifiable parameters from all groups. Figure 3 shows the mode and 95% HPD CrI of the posterior distribution of parameters for groups whose data are able to identify them.

**Fig. 3.**
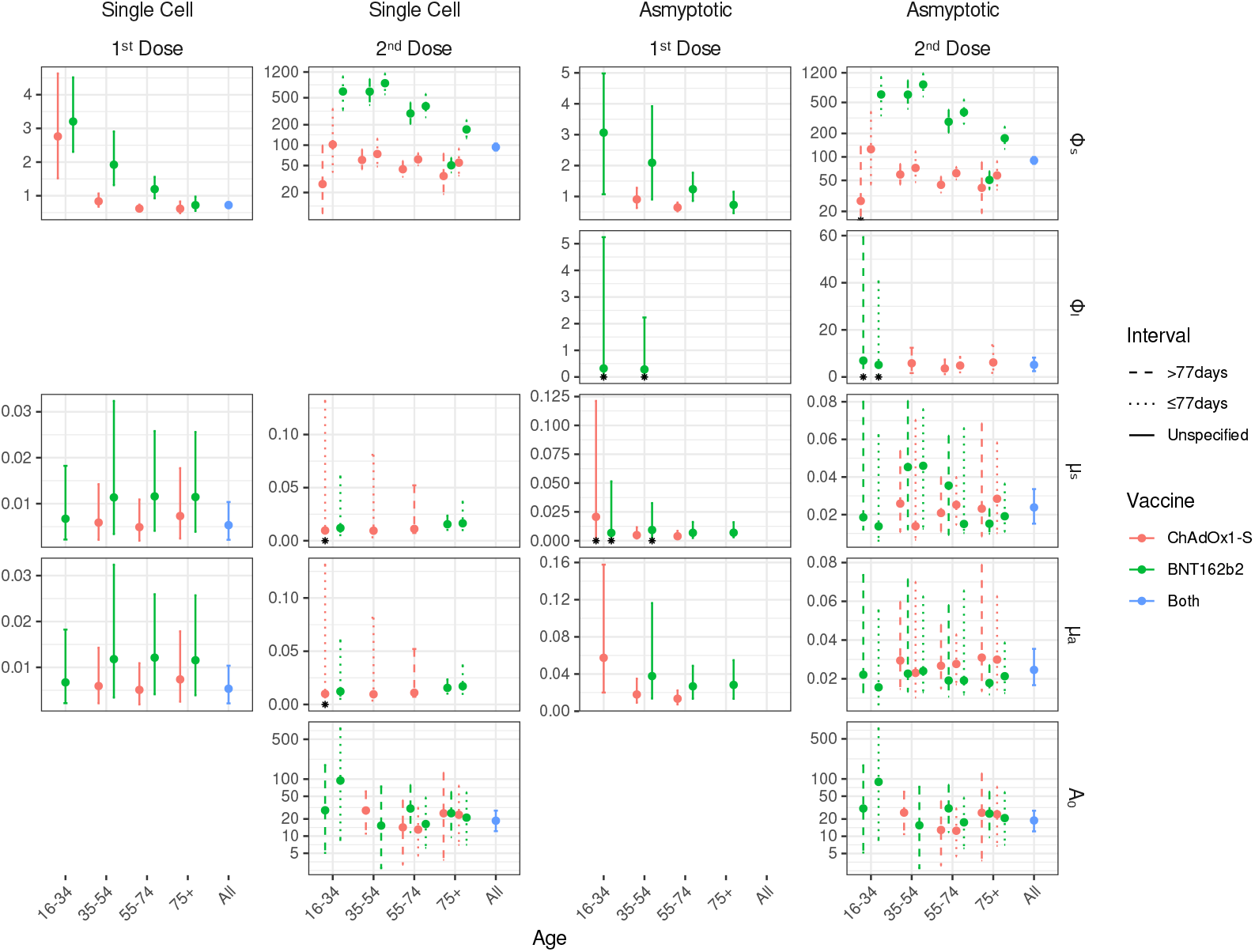
The mode and 95% HDP CrI of the parameter’s posterior distribution for both doses and models, when the parameter is identifiable. The line type is used to distinguish between dosing interval, with dashed and dotted lines used for people who received their second dose *>* 77 days or *≤* 77 days after their first dose respectively, and solid lines are used for groups without a specified dose interval. A dose interval is not specified for first dose groups and when we do not split the data for the dose. Colours are used distinguish between the vaccines. For the second dose only one vaccine is specified for the groups because we only have adequate data for homologously vaccinated people (e.g. someone who received ChAdOx1-S for their second dose also received it for their first dose). Red is used for ChAdOx1-S, green for BNT162b2, and blue for when we do not split the data by vaccine brand. Blue points with blue solid error bars represent the estimates and 95% HDP CrI of parameters fit to all the data on that dose. The black asterisks mark when the lower bound of the parameters 95% HDP CrI is zero.

A common pattern for both models after both doses is that the value of Φ_*s*_ (the antibody production capacity of short-lived plasma cells) decreases with age and is greater for those who received BNT162b2 than those who received ChAdOx1-S. This result explains the higher peak antibody response after vaccination in younger people and people vaccinated BNT162b2 observed in figure 2. Similarly, comparing the estimates of Φ_*s*_ between the first and second doses, we see that both models estimate an increase of 10- to 100- and 70- to 440-fold in Φ_*s*_ when people receive their second dose of ChAdOx1-S and BNT162b2, respectively. These increases explain the higher peaks after two doses observed in figure 2.

A weak distinction between the group estimates of *µ*_*s*_ and *µ*_*a*_ (short-lived plasma cell decay rate and antibody decay rate respectively) in the first dose groups is found by both models. The model fitting suggests people 35 and older who received BNT162b2 have faster rates of plasma cell and antibody decay than those who received ChAdOx1-S. This explains why people vaccinated with one dose of ChAdOx1-S maintain a high antibody level than the same age group who received one dose of BNT162b2, observed in figure 2. However, this pattern is not found after the second dose.

For there to be evidence of a long-term antibody response in a group Φ_*l*_ (the antibody production capacity of long-lived plasma cells) must be greater than zero. Only people 35 and older who received two doses of ChAdOx1-S were found to show strong evidence of a long-term antibody response. For 35-54 year-olds this is only true if they received their doses more than 77 days apart and for people 75 and older this is only true if they received their doses less than 77 days apart. The data collected from 16 – 34 years-olds and 35 – 54 year-olds vaccinated with one BNT162b2, and 16 – 34 years-olds vaccinated with two doses of BNT162b2 was able to identify Φ_*l*_, suggesting these groups could have mounted a long-lived immune response. However, the 95% HDP CrI are [0–5.24], [0–2.23], [0–59.4], and [0–40.6] respectively, all include zero and therefore we cannot be confident long-lived plasma are induced.

Taken together these observations provide quantifiable explanations for observed difference in antibody dynamics between vaccine types and age groups. They show that we only find evidence of a persisting antibody response and thus likely induction of long-lived plasma cells in people vaccinated with two doses of ChAdOx1-S. They also explain greater peak antibody response observed in younger people than older people, people who receive one dose than people who receive two, and people vaccinated with BNT162b2 than people vaccinated with ChAdOx1-S, by more productive short-lived plasma cell (plasmablast) populations.

## Methods and Materials

### Antibody Data

We use SARS-CoV-2 specific antibody data collected from 355,019 routine blood tests between May 2020 and September 2022 collected and validated by the Oxford-Royal College of General Practitioners (RCGP) (see figure 2) (46). This dataset contains vaccination dates and level of total antibody (sum of IgG, IgA, IgM) against the SARS-CoV-2 receptor binding domain (RBD) on the spike (S) protein, measured in units per milliliter (U/ml) at various times after one, two, three or four doses. Due to the sparsity of data after three and four doses, we only use the data after one and two doses.

Previous infection with SARS-CoV-2 can be distinguished from vaccination by seeing whether an individual’s antibodies bind to nucleocapsid (N) protein as well as spike (S) protein. This has been recorded in the data alongside an individual’s vaccine type, number of doses, interval between doses, and age. These factors are used to stratify the population into groups. We assume that the antibody dynamics of the people within a group are the same, therefore individuals’ one-off samples can be interpreted as longitudinal data for the group. As we are investigating the effects of vaccination on antibody levels, we remove anyone who has evidence of previous infection i.e., antibodies that bind to N protein (N positive).

Individuals are categorised by age: 16 – 34 years, 35 – 54 years, 55 – 74 years and 75+ years. We included the BNT162b2 and ChAdOx1-S vaccines and one and two doses and whether the doses were administered less than or equal to 77 days apart or more than 77 days apart (two dose intervals used in the United Kingdom). As we have set four age categories, two vaccine categories, and two dose interval categories, there are 8 possible groups for the first dose and 32 possible groups for the second dose. We have adequate data (i.e. as many time points as parameters for our largest model) for all first dose groups but only 16 of the second dose groups. We do not have adequate data for people who were heterologously vaccinated.

### Mathematical Models

The general model proposed in (18) is the ‘*complete*’ exponential model, that captures the kinetics of short-lived plasma cells *P*_*s*_, long-lived plasma cells *P*_*l*_, and antibodies *A*:

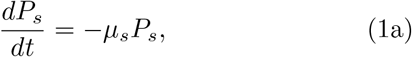

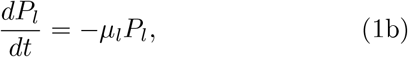

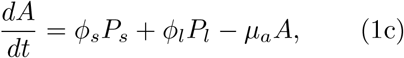

where *µ*_*s*_, *µ*_*l*_ and *µ*_*a*_ denote the decay rate of short-lived plasma cells, long-lived plasma cells, and antibodies respectively, and *ϕ*_*s*_ and *ϕ*_*l*_ denote the rate short- and long-lived plasma cells secretes antibodies, respectively. However, it was found in (18) that assuming *µ*_*l*_ = 0 best describes longitudinal data on hepatitis A specific antibodies with samples taken up to 114 months after vaccination. Hence, we set *µ*_*l*_ = 0, implicitly assuming long-lived plasma cells live forever, and solve the system of equations (1) with the initial conditions 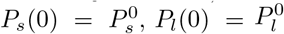, and *A*(0) = *A*_0_ to obtain,

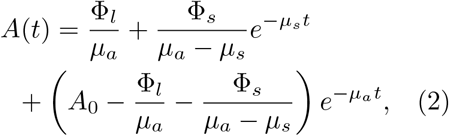

where 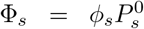 and 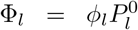. The new parameters Φ_*s*_ and Φ_*l*_ represent the maximum antibody production of the respective plasma cells populations and are defined so that equation (2) is structurally identifiable. We will refer to Φ_*s*_ and Φ_*l*_ as the productive capacity of short-live and long-lived plasma cells, respectively.

We also consider a related model (the single cell model) that assumes long-lived plasma cells are not induced (Φ_*l*_ = 0).

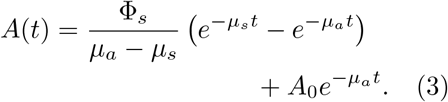

### Model Fitting

We use a Bayesian framework to fit our models (equation (2) and equation (3)) to the data, implemented in R with the rstan package (available at: https://mc-stan.org/). A Markov Chain Monte Carlo (MCMC) algorithm is used to estimate the joint posterior distribution of the parameters.

To obtain a likelihood function we assume that the log of the antibody levels can be modelled as a random variable, *X*(*t*), drawn from a normal distribution with a mean defined our model, *A*(*t, θ*), and a standard deviation *σ*.

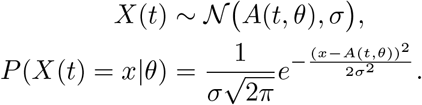

### Initial conditions and prior distributions

When modelling the first dose response we set the initial antibody level, *A*_0_, to zero (as we assume people have not been exposed to SARS-CoV-2) and for the response to the second dose we assume *A*_0_ is log-normally distributed with mean 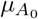 and standard deviation 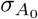. The values of 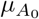 and 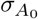 are estimated from the antibody levels of people recorded after their first dose. As the data are cross-sectional, we cannot know what each individual’s antibody level was when they were given their second dose. Hence, we sample the log-transformed serum antibody concentrations after the first dose on the days when the people recorded after their second dose were vaccinated. The mean and standard deviation of this normally distributed sample are used as 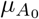 and 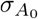 respectively.

For the decay parameters, *µ*_*s*_ and *µ*_*a*_, we use the same log-normal prior for both. A log-normal prior is used because we expect the values of *µ*_*s*_ and *µ*_*a*_ to be small (30; 36; 47; 48; 49; 50; 51; 52; 53), but greater than zero. We calibrate the prior by setting its 2.5% and 97.5% quantiles to 0.01 and 0.1 respectively (equivalent to assuming there is a 95% probability that the true parameter value lies between these values). This reflects the belief that the expected half-lives of antibodies and short-lived plasma cells are in the tens of days (30; 36; 47; 48; 49; 50; 51; 52; 53).

The production capacity parameters, Φ_*s*_ and Φ_*l*_ are more difficult to inform from literature as they are artificial. However, Φ_*l*_ can still be informed through assumptions about the long-term dynamics of the model. The resting antibody level at *t*_*∞*_ is predicted by the asymptotic model (equation 2) to be 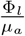, so Φ_*l*_ will be equal to the product of *µ*_*a*_ and the resting antibody level, which we denote lAb_*∞*_. We have assumed that both the antibody data and *µ*_*a*_ are log-normally distributed. The product of two log-normal distributions is log-normally distributed. If we say the mean and standard deviation of the prior for *µ*_*a*_ is *µ*_decay_ and *σ*_decay_ respectively, and the mean and standard deviation of the resting antibody level is *µ*_*∞*_ and *σ*_*∞*_ respectively, the prior for Φ_*l*_ will have the mean and standard deviation 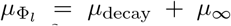 and 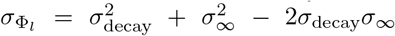. We calculate *µ*_*∞*_ and *σ*_*∞*_ through assuming the 2.5% and 97.5% quantiles of the distribution. After the first dose we assume the 2.5% and 97.5% quantiles of the log-transformed resting antibody level will be 2 and 7 respectively and after the second dose the 3 and 8, respectively. These quantiles are informed by the data we have on earlier time points, therefore not violating the principle of prior belief being ignorant to data, as we do not have data for long-time.

The final parameter is the production capacity of short-lived plasma cells, Φ_*s*_. There is not the same intuitive interpretation for Φ_*s*_ in the dynamics of the model as there is for Φ_*l*_ so we use a truncated normal distribution with the 2.5% and 97.5% quantiles 1 and 5 respectively for the first dose and 50 to 1000 respectively for the second dose. These quantiles were calibrated through prior predictive checks.

To summarise, the prior distributions are,

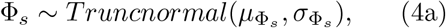

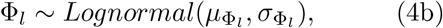

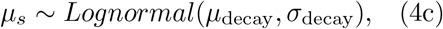

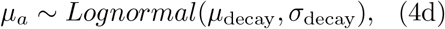

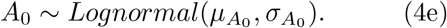

### Parameter Identifiability

We use the STRIKE GOLDD Matlab package (54) (available at https://github.com/afvillaverde/strike-goldd) to determine whether our models are structurally identifiable. We use STRIKE GOLDD because it considers initial condition as parameters and has tools for reparameterization in the case of structural non-identifiability.

The true value of a structural identifiable parameter is theoretically possible to find, given noise-free data. It may not be possible to determine the true value of a structurally identifiable parameter when fitting to noisy or sparse data. Practical identifiability, defined as the ability to define finite bounds around an estimate (55), determines whether the data contain enough information to estimate the parameter value with confidence.

An established method of defining confidence intervals is with the profile likelihood method (55), by assuming values of the parameter and maximising the likelihood function with respect to the remaining parameters (the *‘nuisance’* parameters) (55; 56; 57). The simplest implementation of maximisation uses maximum likelihood estimation (MLE) and gradient decent methods (55; 57). Though the methodology of the profile likelihood method is well defined it can be fragile when likelihood spaces are complex and difficult to navigate by gradient decent.

To mitigate the fragility of the likelihood profile method we use a Monte Carlo Markov Chain (MCMC) algorithm to explore the parameter space more completely and generate posterior distributions of parameter values. We determine a parameter to be practically identifiable if the 95% highest posterior density credible interval (HPD CrI) of the posterior distribution is constrained to a single biologically meaningful set and is not sensitive to prior assumptions. We say the parameter is not identifiable when the 95% HPD CrI is made up of multiple separate regions because this implies the data do not contain enough information to constrain the parameter estimate around the true value. Similarly, if the 95% HPD CrI is sensitive to prior assumptions this implies the data do not contain enough information to inform the parameter.

### Sensitivity analysis

We conduct the sensitivity analysis by expanding the quantiles of the prior distributions of Φ_*s*_, *µ*_*s*_, *µ*_*a*_, lAb_*∞*_, and doubling the standard deviation calculated for a particular group, shown in table 1. To determine whether a 95% HPD CrI is sensitive to the priors we check two things: whether the inclusion of zero in the HPD CrI has changed, and whether the modes of both posteriors are included within the HPD CrI of both posteriors. If the inclusion of zero in HPD CrI changes, so does the conclusion about whether the effect of the parameter can be detected in the data. This is a fundamental change, and we say the posteriors are too sensitive to the prior in this case. Checking if the modes of the posteriors are included within the HPD CrI of the other posterior is to test for agreement in the estimates. If there is agreement, we say the posterior is not too sensitive to the priors.

**Table 1.** Prior distributions for the model parameters. For Φ_*s*_, *µ*_*s*_, *µ*_*a*_, and lAb_*∞*_ we assume the 2.5% and 97.5% quantiles of the prior distributions. For *A*_0_ the assumptions are the mean and standard deviation. Values are not given for the *A*_0_ assumptions as they vary between the groups.

| Dose | Baseline Prior Assumptions |  |  |  |  |
| --- | --- | --- | --- | --- | --- |
| | $\Phi_s$ | $\mu_s$ | $\mu_a$ | $\text{lAb}_\infty$ | $A_0$ |
| 1<sup>st</sup> | [1, 5] | [0.01, 0.1] | [0.01, 0.1] | [2, 7] | - |
| 2<sup>nd</sup> | [50, 1000] | [0.01, 0.1] | [0.01, 0.1] | [3, 8] | $\mu_{A_0}, \sigma_{A_0}$ |
| Dose | Sensitivity Analysis Prior Assumptions |  |  |  |  |
| | $\Phi_s$ | $\mu_s$ | $\mu_a$ | $\text{lAb}_\infty$ | $A_0$ |
| 1<sup>st</sup> | [0.5, 10] | [0.005, 0.1] | [0.005, 0.1] | [1, 8] | - |
| 2<sup>nd</sup> | [50, 2000] | [0.005, 0.1] | [0.005, 0.1] | [2, 9] | $\mu_{A_0}, 2\sigma_{A_0}$ |

**Table 2.** The vaccines schedule, age, size, best fitting model, parameter estimates and 95% HDP CrI of the groups whose data could identify a model. ‘*Pfz* ‘ and ‘*Az* ‘ are short hand for BNT162b2 and ChAdOx1-S respectively. ‘*SC* ‘ and ‘*Asym*’ are short hand for the single cell and asymptotic models respectively. The parameters Φ_*s*_, Φ_*l*_, *µ*_*s*_, *µ*_*a*_, and *A*_0_ are the short-lived plasma cell productive capacity, the long-lived plasma cell productive capacity, the short-lived plasma cell decay rate, the antibody decay rate, and the initial level of antibody respectively.

| Vaccine<br>1<sup>st</sup> | 2<sup>nd</sup> | Age | Dose<br>Interval | Sample<br>Size | Model | $\Phi_s$ | $\Phi_l$ | Parameter Estimates | | |
| --- | --- | --- | --- | --- | --- | --- | --- | --- | --- | --- |
| | | | | | | | | $\mu_s$ | $\mu_a$ | $A_0$ |
| Az | - | 35 – 54 | - | 352 | SC | 0.835<br>[0.673 – 1.07] | - | 0.00588<br>[0.00214 – 0.0142] | 0.00593<br>[0.00216 – 0.0143] | - |
| Az | - | 55 – 74 | - | 870 | SC | 0.621<br>[0.535 – 0.738] | - | 0.00490<br>[0.00193 – 0.0109] | 0.00509<br>[0.00191 – 0.0109] | - |
| Az | - | 75+ | - | 300 | SC | 0.615<br>[0.481 – 0.831] | - | 0.00731<br>[0.00241 – 0.0177] | 0.00737<br>[0.00251 – 0.0179] | - |
| Pfz | - | 16 – 34 | - | 92 | SC | 3.20<br>[2.30 – 4.52] | - | 0.00672<br>[0.00219 – 0.0182] | 0.00673<br>[0.00219 – 0.0182] | - |
| Pfz | - | 35 – 54 | - | 182 | SC | 1.92<br>[1.31 – 2.91] | - | 0.0114<br>[0.00337 – 0.0324] | 0.0118<br>[0.00341 – 0.0324] | - |
| Pfz | - | 55 – 74 | - | 532 | SC | 1.20<br>[0.923 – 1.56] | - | 0.116<br>[0.00409 – 0.0258] | 0.0121<br>[0.00413 – 0.0260] | - |
| Pfz | - | 75+ | - | 547 | SC | 0.722<br>[0.551 – 0.976] | - | 0.0114<br>[0.00388 – 0.0256] | 0.0116<br>[0.00391 – 0.0257] | - |
| Az | Az | 35 – 54 | >77days | 453 | Asym | 59.5<br>[44.2 – 81.3] | 5.82<br>[1.67 – 12.4] | 0.0259<br>[0.0107 – 0.0539] | 0.0294<br>[0.0147 – 0.0598] | 25.8<br>[10.9 – 59.7] |
| Az | Az | 55 – 74 | ≤77days | 967 | Asym | 61.8<br>[51.5 – 74.5] | 4.85<br>[1.96 – 8.56] | 0.0252<br>[0.0146 – 0.0400] | 0.0276<br>[0.0171 – 0.0429] | 12.5<br>[4.45 – 30.7] |
| Az | Az | 55 – 74 | >77days | 695 | Asym | 43.8<br>[34.9 – 55.9] | 3.60<br>[1.20 – 7.43] | 0.0209<br>[0.0109 – 0.0400] | 0.0267<br>[0.0149 – 0.0479] | 12.9<br>[3.07 – 41.1] |
| Az | Az | 75+ | ≤77days | 285 | Asym | 57.9<br>[37.9 – 88.3] | 6.16<br>[1.74 – 13.5] | 0.0284<br>[0.0102 – 0.0584] | 0.0299<br>[0.0144 – 0.0627] | 24.2<br>[6.79 – 73.2] |
| Pfz | Pfz | 16 – 34 | ≤77days | 25 | Asym | 632<br>[341 – 1060] | 5.15<br>[0 – 40.6] | 0.0138<br>[0.00612 – 0.0622] | 0.0156<br>[0.00674 – 0.0553] | 88.9<br>[8.64 – 761] |
| Pfz | Pfz | 75+ | ≤77days | 430 | SC | 170<br>[126 – 234] | - | 0.0164<br>[0.0100 – 0.0366] | 0.0171<br>[0.00992 – 0.0367] | 21.1<br>[7.05 – 58.8] |
| Pfz | Pfz | 75+ | >77days | 661 | SC | 50.4<br>[39.5–65.0] | - | 0.0155<br>[0.0102–0.0236] | 0.0156<br>[0.0102–0.0235] | 25.2<br>[9.55–59.8] |

## Discussion

We have proposed and analysed two mechanistic and identifiable models of SARS-CoV-2 specific antibody dynamics that link cross-sectional seroprevalence data to within-host antibody dynamics. Our models relate peak antibody levels to productive capacity of short-lived plasma cells, explaining the observation that individuals who received BNT162b2 have higher peak levels than those that received ChAdOx1-S (7), as well as a higher peak observed in younger people than older people (4; 1; 2; 3). Persistent antibody levels observed in people who receive two doses of ChAdOx1-S are presumed to be explained by the induction of long-lived plasma cells. We found no effect of age on the apparent production capacity of long-lived plasma cells.

The inability of mRNA COVID-19 vaccines to elicit long-lived plasma cells after one and two doses has been discussed previously (58). However, long-lived plasma cells in bone marrow, and improved immunogenicity, have been found in people vaccinated with three and four doses of mRNA COVID-19 vaccines (59; 60), although this may be due to infection.

Previous mathematical models of SARS-CoV-2 antibody responses were phenomenologically rather than biologically motivated, often due to data limitations (11; 12; 13; 14; 15; 16). These models considered a constant rate of antibody decay and do not account for the production of antibodies by plasma cells. Therefore, they cannot inform on the mechanisms of antibody maintenance and have limited power to predict future antibody levels. Our single cell and asymptotic models consider two and three time-frames respectively, allowing them to distinguish separate phases of decay naturally. When models only consider a single phase of decay (11; 12; 13; 14; 15; 16) model calibration is forced to compromise between short-term forces (short-lived plasma cells, antibody decay) and long-term forces (long-lived plasma cells), so parameter estimates and predicted dynamics can be difficult to interpret.

Previous modelling estimated the half-life of antibodies to be hundreds of days (11; 12; 13) with wide confidence intervals (11). In contrast, our model explicitly captures the kinetics of antibody production and our point estimates of the half-life of antibodies (23 – 116 days) are consistent with experimental findings (47; 49; 50; 51; 52; 36) and previous estimates (15; 16). Our improved estimates are due to our approach’s ability to overcome the restrictions of single-phase models with a description of antibody dynamics that considers their production. Single-phase models overestimate the half-life of antibodies to compensate for the lack of antibody renewal. Half-life estimates of from single-phase models can depend on how many doses a person received (16). The change in estimates following booster doses will likely be due to the effects of memory B cells that become dominant in the humoral response, and the induction of long-lived plasma cells, effects our model captures and overcomes.

A limitation of our models is the assumption that the initial population of plasma cells is the full population that decays over time. In actuality, after vaccination, antigen will be produced within the host’s cells (61), presented to naive and memory B cells within the lymph nodes by follicular dendritic cells, which then through various routes proliferate and differentiate into plasma cells (62). A more accurate model could include the initial expansion of the plasma cell population. Such a model would fit the initial antibody response better and give information on differences in plasma cell generation post vaccination between population groups. The improved fitting would be most pronounced for responses to the first dose as that is when we would not expect people to have SARS-CoV-2 specific plasma cells prior to vaccination. However, such a model would require data of other immune factors such as germinal center B cells, which are not easy to obtain. Another limitation is the length of time over which the data were collected. Data that continued further after each dose would help clarify long-time dynamics and help validate our predictions. Due to identifiability issues, it is not possible for our models to distinguish the plasma cell number and rate of antibody production from each other. So, we cannot determine what combination of these two factors is responsible for variation in the overall production of antibodies between groups.

Our models suggest that while mRNA vaccines induce larger short term responses which may have advantages for rapid protection, the adenoviral vector vaccines may have advantages in eliciting persistent serum antibodies against SARS-CoV-2, presumably through induction of long-lived plasma cells. Vaccinating people who have only received mRNA vaccines with adenoviral vector vaccines, with priority given to older people, may achieve persistently high antibodies in all age groups. Clinical studies into the effectiveness of BNT162b2 and ChAdOx1-S against infection with SARS-CoV-2 and hospitalisation with COVID-19 find that BNT162b2 and ChAdOx1-S are comparable after one dose, but BNT162b2 is more effective after two doses (63; 64; 65; 66; 67; 68; 69). However, comparisons of BNT162b2 and ChAdOx1-S vaccine effectiveness are usually short term, at most up to 30 weeks, whereas our models cover longer timescales. Further, we cannot be certain whether heterologous vaccination will elicit the same humoral response and effectiveness as homologous vaccination. Clinical studies have found that heterologous vaccination is more effective than homologous vaccination at protecting against infection (70; 71; 72; 73). However, these findings are for adenoviral vector-primed mRNA-boosted individuals while we are discussing boosting mRNA-primed individuals with adenoviral vector vaccine. It has been found that binding and neutralizing antibodies wane less in individuals primed with mRNA vaccine and boosted with adenoviral vaccine than in individuals vaccinated with other heterologous vaccination regimes (74; 75), supporting mRNA priming and adenoviral vector boosting to achieve persistently high antibody levels.

## Conclusion

Routinely-collected seroprevalence data are valuable for characterising within-host mechanisms of antibody production and persistence. Extended sampling and linking seroprevalence data to outcomes would allow for powerful conclusions on the relationship between humoral kinetics and protection against disease. With added data on other immune factors, such as serum memory B cell and plasmablasts, mathematical modelling can give more accurate descriptions of the complex kinetics of the humoral response. Closer interaction between mathematical modelers and immunologists on data collection and model structure will equip interdisciplinary groups with the necessary information to describe population immune responses more accurately, benefiting the fields of immunology and epidemiology. With the current data we can make projections of antibody responses into the future and our results suggest older people who have not received adenoviral vector vaccine may derive longer lasting protection from a booster dose of this vaccine type than from additional mRNA vaccination.

## Data Availability

The United Kingdom Health and Security Agency (UKHSA) dataset can be accessed by researchers; approval is on a project-by-project basis (https://orchid.phc.ox.ac.uk/index.php/orchid-data/).

## Acknowledgments

The authors would like to thank Heather Whitaker and the Oxford-Royal College of General Practitioners Research and Surveillance Centre (Oxford-RCGP RSC) as well as UKHSA for sampling, testing, and data collection. DS is supported by Engineering and Physical Sciences Research council (EPSRC) PhD studentship (grant number EP/W524414/1). We would like to acknowledge the help and support of the JUNIPER partnership (MRC grant no MR/X018598/1) which EBP, LD and DS are affiliated with. EBP receives funding from the NIHR Health Protection Research Unit in Behavioural Science and Evaluation at University of Bristol. LD is further supported by Pfizer through investigator-led grants on respiratory tract infections.

## References

[1] Weisberg, S. P. et al. Distinct antibody responses to SARS-CoV-2 in children and adults across the COVID-19 clinical spectrum. Nature Immunology 22, 25—31 (2021).

[2] Dowell, A. C. et al. Children develop robust and sustained cross-reactive spike-specific immune responses to SARS-CoV-2 infection. Nature Immunology 23, 40—49 (2022).

[3] Collier, D. A. et al. Age-related immune response heterogeneity to SARS-CoV-2 vaccine BNT162b2. Nature 596, 417–422 (2021).

[4] Wei, J. et al. Antibody responses to SARS-CoV-2 vaccines in 45,965 adults from the general population of the united kingdom. Nature Microbiology 6, 1140–1149 (2021).

[5] Evans, J. P. et al. Neutralizing antibody responses elicited by SARS-CoV-2 mRNA vaccination wane over time and are boosted by break-through infection. Science Transla-tional Medicine 14 (2022).

[6] Qi, H. et al. The humoral response and antibodies against SARS-CoV-2 infection. Nature Immunology. 23, 1008–1020 (2022).

[7] Rose, R. et al. Humoral immune response after different SARS-CoV-2 vaccination regimens. BMC Medicine 20 (2022).

[8] Goel, R. R. et al. Distinct antibody and memory B cell responses in SARS-CoV-2 naïve and recovered individuals after mRNA vaccination Science Immunology, eabi6950 (2021)

[9] Geers, D. et al. SARS-CoV-2 variants of concern partially escape humoral but not T cell responses in COVID-19 convalesSscent donors and vaccine recipients. Science Immunology 6, eabj1750 (2021)

[10] Becker, M. et al. Immune response to SARS-CoV-2 variants of concern England in vaccinated individuals. Nature Communications 12 (2021).

[11] Whitcombe, A. L. et al. Comprehensive analysis of SARS-CoV-2 antibody dynamics in New Zealand. Clinical & Translational Immunology 10 (2021).

[12] Dorigatti, I. et al. SARS-CoV-2 antibody dynamics and transmission from community-wide serological testing in the Italian municipality of Vo’. Nature Communications 12 (2021).

[13] Chen, Y. et al. Immune recall improves antibody durability and breadth to SARS-CoV-2 variants. Science Immunology 7 (2022).

[14] Achiron, A. et al. SARS-CoV-2 antibody dynamics and b-cell memory response over time in COVID-19 convalescent subjects. Clinical Microbiology and Infection 27, 1349.e1–1349.e6 (2021).

[15] Lumley, S. F. et al. The duration, dynamics, and determinants of severe acute respiratory syndrome coronavirus 2 (SARS-CoV-2) anti-body responses in individual health-care workers. Clinical Infectious Diseases 73, e699–e709 (2021).

[16] Yorsaeng, R. et al. SARS-CoV-2 antibody dynamics after COVID-19 vaccination and infection: A real-world cross-sectional analysis. Vaccines 11, 1184 (2023).

[17] Manz, R. A. et al. Maintenance of serum antibody levels. Annual Reviews Immunology 23, 367–386 (2005).

[18] Andraud, M. et al. Living on three time scales: The dynamics of plasma cell and antibody populations illustrated for hepatitis A virus. PLoS Computational Biology 8, e1002418 (2012).

[19] Fraser, C. et al. Modeling the long-term antibody response of a human papillomavirus (HPV) virus-like particle (VLP) type 16 prophylactic vaccine. Vaccine 25, 4324–4333 (2007).

[20] Benner, R., Hijmans, W. & Haaijman, J. J. The bone marrow: the major source of serum immunoglobulins, but still a neglected site of antibody formation. Clinical and Experimental Immunology 46, 1–8 (1981).

[21] Hyland, L., Sangster, M., Sealy, R. & Coleclough, C. Respiratory virus infection of mice provokes a permanent humoral immune response. Journal of Virology 68, 6083–6086 (1994).

[22] Hill, S. W. Distribution of plaque-forming cells in the mouse for a protein antigen. Evidence for highly active parathymic lymph nodes following intraperitoneal injection of hen lysozyme. Immunology 30, 895– 906 (1976).

[23] Slifka, M. K., Matloubian, M. & Ahmed, R. Bone marrow is a major site of long-term antibody production after acute viral infection. Journal of Virology 69, 1895– 1902 (1995).

[24] Earle, K. A. et al. Evidence for antibody as a protective correlate for COVID-19 vaccines. Vaccine 39, 4423–4428 (2021).

[25] Fong, Y. et al. Immune correlates analysis of the PREVENT-19 COVID-19 vaccine efficacy clinical trial. Nature Communications 14, 331 (2023).

[26] Woolsey, C. et al. Establishment of an African green monkey model for COVID-19 and protection against re-infection. Nature Immunology 22, 86–98 (2021).

[27] Mink, S. et al. Evaluation of SARS-CoV-2 antibody levels on hospital admission as a correlate of protection against mortality. Journal of Internal Medicine 293, 694–703 (2023).

[28] Amanna, I. J. & Slifka, M. K. Mechanisms that determine plasma cell lifespan and the duration of humoral immunity. Immunological Reviews 236, 125–138 (2010).

[29] Amanna, I. J., Carlson, N. E. & Slifka, M. K. Duration of humoral immunity to common viral and vaccine antigens. New England Journal of Medicine 357, 1903–1915 (2007).

[30] Slifka, M. & Ahmed, R. Long-term humoral immunity against viruses: revisiting the issue of plasma cell longevity. Trends in Microbiology 4, 394–400 (1996).

[31] Slifka, M. K. Immunological memory to viral infection. Current Opinion in Immunology 16, 443–450 (2004).

[32] MacLennan, I. C. M. Germinal centers. Annual Review of Immunology 12, 117–139 (1994).

[33] Liu, Y.-J., Grouard, G., de Bouteiller, O. & Banchereau, J. Follicular dendritic cells and germinal centers. International Review of Cytology 166 139–179 (1996).

[34] Tew, J. G., Kosco, M. H., Burton, G. F. & Szakal, A. K. Follicular dendritic cells as accessory cells. Immunological Reviews 117, 185–211 (1990).

[35] Tew, J. G. & Mandel, T. E., Prolonged antigen half-life in the lym-phoid follicles of specifically immu-nized mice. Immunology 37, 69–76 (1997).

[36] Hammarlund, E. et al. Plasma cell survival in the absence of B cell memory. Nature Communications 8 (2017).

[37] Manz, R. A., Thiel, A. & Radbruch, A. Lifetime of plasma cells in the bone marrow. Nature 388, 133–134 (1997).

[38] Ahuja, A., Anderson, S. M., Khalil, A. & Shlomchik, M. J. Maintenance of the plasma cell pool is independent of memory B cells. Proceedings of the National Academy of Sciences 105, 4802–4807 (2008).

[39] DiLillo, D. J. et al. Maintenance of long-lived plasma cells and serological memory despite mature and memory B cell depletion during CD20 immunotherapy in mice. The Journal of Immunology 180, 361– 371 (2008).

[40] Slifka, M. K., Antia, R., Whitmire, J. K. & Ahmed, R. Humoral immunity due to long-lived plasma cells. Immunity 8, 363–372 (1998).

[41] Radbruch, A. et al. Competence and competition: the challenge of becoming a long-lived plasma cell. Nature Reviews Immunology 6, 741–750 (2006).

[42] Garrido, C. et al. SARS-CoV-2 vaccines elicit durable immune responses in infant rhesus macaques. Science Immunology 6, eabj3684 (2021).

[43] Tunali, E. & Tarn, T.-J. New results for identifiability of nonlinear systems. IEEE Transactions on Automatic Control 32, 146–154 (1987).

[44] Ljung, L. & Glad, T. On global identifiability for arbitrary model parametrizations. Automatica 30, 265–276 (1994).

[45] Xia, X. & Moog, C. Identifiability of nonlinear systems with application to HIV/AIDS models. IEEE Transactions on Automatic Control 48, 330–336 (2003).

[46] Whitaker, H. et al. Sociodemographic disparities in COVID-19 seroprevalence across England in the Oxford RCGP primary care sentinel network Journal of Infection 84, 814–824 (2022)

[47] Morell, A., Terry, W. D. & Waldmann, T. A. Metabolic properties of IgG subclasses in man. Journal of Clinical Investigation 49, 673–680 (1970).

[48] Spiegelberg, H. L. & Fishkin, B. G. The catabolism of human g immunoglobulins of different heavy chain subclasses. 3. the catabolism of heavy chain disease proteins and of fc fragments of myeloma proteins. Clinical and Experimental Immunology 10, 599–607 (1972).

[49] Tam, S. H., McCarthy, S. G., Brosnan, K., Goldberg, K. M. & Scallon, B. J. Correlations between pharmacokinetics of IgG antibodies in primates vs. FcRn-transgenic mice reveal a rodent model with predictive capabilities. mAbs 5, 397–405 (2013).

[50] Robbie, G. J. et al. A novel investigational fc-modified humanized monoclonal antibody, motavizumab-YTE, has an extended half-life in healthy adults. Antimicrobial Agents and Chemotherapy 57, 6147–6153 (2013).

[51] Hopkins, R. J. et al. Safety and pharmacokinetic evaluation of intravenous vaccinia immune globulin in healthy volunteers. Clinical Infectious Diseases 39, 759–766 (2004).

[52] Adner, N. et al. Pharmacokinetics of human tick-borne encephalitis virus antibody levels after injection with human tick-borne encephalitis immunoglobulin, solvent/detergent treated, FSME-BULIN s/d in healthy volunteers. Scandinavian Journal of Infectious Diseases 33, 843–847 (2001).

[53] Goulet, D. R. et al. Toward a combinatorial approach for the prediction of IgG half-life and clearance. Drug Metabolism and Disposition 46, 1900–1907 (2018).

[54] Villaverde, A. F., Barreiro, A. & Papachristodoulou, A. Structural identifiability of dynamic systems biology models. PLOS Computational Biology 12, e1005153 (2016).

[55] Raue, A. et al. Structural and practical identifiability analysis of partially observed dynamical models by exploiting the profile likelihood. Bioinformatics 25, 1923–1929 (2009).

[56] Murphy, S. A. & Vaart, A. W. V. D. On profile likelihood. Journal of the American Statistical Association 95, 449–465 (2000).

[57] Meeker, W. Q. & Escobar, L. A. Teaching about approximate confidence regions based on maximum likelihood estimation. The American Statistician 49, 48 (1995).

[58] Giannotta, G. & Giannotta, N. mRNA COVID-19 vaccines and long-lived plasma cells: A complicated relationship. Vaccines 9, 1503 (2021).

[59] Schulz, A. R. et al. SARS-CoV-2 specific plasma cells acquire long-lived phenotypes in human bone marrow EBioMedicine 95, (2023)

[60] Lustig, Y. et al. Superior immunogenicity and effectiveness of the third compared to the second BNT162b2 vaccine dose. Nature Immunology 23, 940—946 (2022).

[61] Gebre, M. S. et al. Novel approaches for vaccine development. Cell 184, 1589–1603 (2021).

[62] Murphy, K. & Weaver, C. Janeway’s Immunobiology. 10, Garland science (2021)

[63] Flacco, M. E. et al. Interim estimates of COVID-19 vaccine effectiveness in a mass vaccination setting: data from an Italian Province. Vaccines 9, 628 (2021).

[64] Bernal, J. L. et al. Effectiveness of the Pfizer-BioNTech and Oxford-AstraZeneca vaccines on COVID-19 related symptoms, hospital admissions, and mortality in older adults in England: test negative case-control study. BMJ 373, (2021).

[65] Bernal, J. L. et al. Effectiveness of COVID-19 Vaccines against the B.1.617.2 (Delta) Variant. NEJM 385, 585–594 (2021).

[66] Paris, C. et al. Effectiveness of mRNA-BNT162b2, mRNA-1273, and ChAdOx1 nCoV-19 vaccines against COVID-19 in healthcare workers: an observational study using surveillance data. Clinical Microbiology and Infection 27, 1699–e5 (2021).

[67] Hyams, C. et al. Effectiveness of BNT162b2 and ChAdOx1 nCoV-19 COVID-19 vaccination at preventing hospitalisations in people aged at least 80 years: a test-negative, case-control study. The Lancet Infectious Diseases 21, 1539–1548 (2021).

[68] Horne, E. M. et al. Waning effectiveness of BNT162b2 and ChAdOx1 COVID-19 vaccines over six months since second dose: OpenSAFELY cohort study using linked electronic health records. BMJ 378, (2022).

[69] Xie, J. et al. Comparative effectiveness of the BNT162b2 and ChAdOx1 vaccines against COVID-19 in people over 50. Nature Communications 13, (2022).

[70] Nordstörm, P., Ballin, M. & Nordström, A. Effectiveness of heterologous ChAdOx1 nCoV-19 and mRNA prime-boost vaccination against symptomatic COVID-19 infection in Sweden: A nationwide cohort study. The Lancet Regional Health–Europe 11, (2021).

[71] Pozzetto, B. et al. Immuno-genicity and efficacy of heterolo-gous ChAdOx1–BNT162b2 vaccination. Nature 600, 701–706 (2021).

[72] Hermossilla, E. et al. Comparative effectiveness and safety of homologous two-dose ChAdOx1 versus heterologous vaccination with ChAdOx1 and BNT162b2. Nature Communications 13, (2022).

[73] Marra, A. R. et al. Effectiveness of Heterologous Coronavirus Disease 2019 (COVID-19) Vaccine Booster Dosing in Brazilian Health-care Workers, 2021. Clinical Infectious Diseases 76, e360.#x2014;e366 (2022).

[74] Atmar, R. L. et al. Homologous and Heterologous COVID-19 Booster Vaccinations. NEJM 386, 1046–1057 (2022).

[75] Tan, C. S. et al. Durability of Heterologous and Homologous COVID-19 Vaccine Boosts. JAMA Network Open 5, e2226335 (2022).

